# Effect of large-scale testing platform in prevention and control of the COVID-19 pandemic: an empirical study with a novel numerical model

**DOI:** 10.1101/2020.03.15.20036624

**Authors:** Qing Xie, Jing Wang, Jianling You, Shida Zhu, Rui Zhou, Zhijian Tian, Hao Wu, Yang Lin, Wei Chen, Lan Xiao, Xin Jin, Jianjuan Li, Jie Dong, Honglong Wu, Wei Zhang, Jing Li, Xun Xu, Ye Yin, Feng Mu, Weijun Chen, Wang Jian

## Abstract

**Background:** China adopted an unprecedented province-scale quarantine since January 23^rd^ 2020, after the novel coronavirus (COVID-19) broke out in Wuhan in December 2019. Responding to the challenge of limited testing capacity, large-scale standardized and fully-automated laboratory (Huo-Yan) was built as an ad-hoc measure. There was so far no empirical data or mathematical model to reveal the impact of the testing capacity improvement since the quarantine.

**Methods:** We integrated public data released by the Health Commission of Hubei Province and Huo-Yan Laboratory testing data into a novel differential model with non-linear transfer coefficients and competitive compartments, to evaluate the trends of suspected cases under different nucleic acid testing capacities.

**Results:** Without the establishment of Huo-Yan, the suspected cases would increased by 47% to 33,700, the corresponding cost of the quarantine would be doubled, and the turning point of the increment of suspected cases and the achievement of “daily settlement” (all daily new discovered suspected cases were diagnosed according to the nucleic acid testing results) would be delayed for a whole week and 11 days. If the Huo-Yan Laboratory ran at its full capacity, the number of suspected cases would decrease at least a week earlier, the peak of suspected cases would be reduced by at least 44% and the quarantine cost could be reduced by more than 72%. Ideally, if a daily testing capacity of 10,500 could achieved immediately after the Hubei lockdown, “daily settlement” for all suspected cases would be achieved immediately.

**Conclusions:** Large-scale and standardized clinical testing platform with nucleic acid testing, high-throughput sequencing and immunoprotein assessment capabilities need to be implemented simultaneously in order to maximize the effect of quarantine and minimize the duration and cost. Such infrastructure like Huo-Yan, is of great significance for the early prevention and control of infectious diseases for both common times and emergencies.

## Introduction

To cope with the outbreak of the coronavirus related disease (COVID-19) in Wuhan since December 2019, the unprecedented province-scale quarantine since January 23^rd^ 2020 was adopted to prevent the virus from spreading ^[1,2]^. The numerical simulation of Yang *et al*. ^[1]^ quantitatively explained the effectiveness of the series of unprecedented measures taken by the Chinese government, such as extended the Spring Festival holiday, encouraged people to self-quarantined and delayed the resumption of work and school, which successfully reduced the population movement and thereby the virus transmission.

The clinical testing method plays irreplaceable role in identifying the infected, cutting off the transmission, and protecting the susceptible. The qRT-PCR based nucleic acid testing is regarded as one of the gold standards for the detection of coronavirus related disease (COVID-19). From January 3^rd^, the Chinese Center for Disease Control and Prevention (CCDCP) began to distribute nucleic acid testing kits to hospitals and medical institutions, and carried out testings according to their own capabilities. However, the large-scale and standardization nucleic acid testing has always been a problem that troubles the entire disease control system, including the CCDCP, hospitals and clinical laboratories. Quality control of the sampling procedure, equipments, testing kits and processes lack consistency amongst 97 institutions in Hubei Province and more than 40 institutions in Wuhan, making it hard to centralize and scale-up the testings, deliver the results and admit the infected on a timely manner. The above mentioned technical issues of the clinical testing lead to the controversy about the effectiveness of the nucleic acid testing by the doctors, experts and officials, which converted into the social panic. On February 4^th^, the fifth edition of the diagnosis and treatment plan for the novel coronavirus disease even adapted the imaging features of pneumonia (by CT-scanning) as the diagnosis standard of COVID-19 in Hubei Province published by the National Health Commission ^[3]^.

Though the province-scale quarantine is unprecedented, however the number of suspected infections kept increasing due to a series issues with regard to the nucleic acid testing, which leading to serious delay of both diagnosis and hospital admission. To cope with that, the Wuhan government made another key strategic decision to build an emergent clinical virus testing infrastructure on Jan 29^th^, i.e. the Huo-Yan Laboratory, with a testing capacity over 10,000 per day (Figure 1). Huo-Yan was expanded into a site of 2,000 m^2^ within a week from an existing laboratory that continuously delivers testing results. Since Huo-Yan put into use on Feb 5^th^, its testing capacity kept stably increasing due to the automated nucleic acid extraction device and optimization of procedure. Then Huo-Yan have achieved 14,000 testing capacity per day on Feb 9^th^ along with the original site and exceeded 20,000 testing capacity per day on Mar 1^st^. And finally substantially contributed to achieve the “daily settlement” (no suspected cases each day) raised by Hubei Provincial government starting by Feb 16^th^. On Feb 19^th^, the sixth edition of the diagnosis and treatment plan for the novel coronavirus disease ^[3]^ recalled the practice of using imaging features of pneumonia for the diagnosis of COVID-19 in Hubei Province.

**Figure 1.**
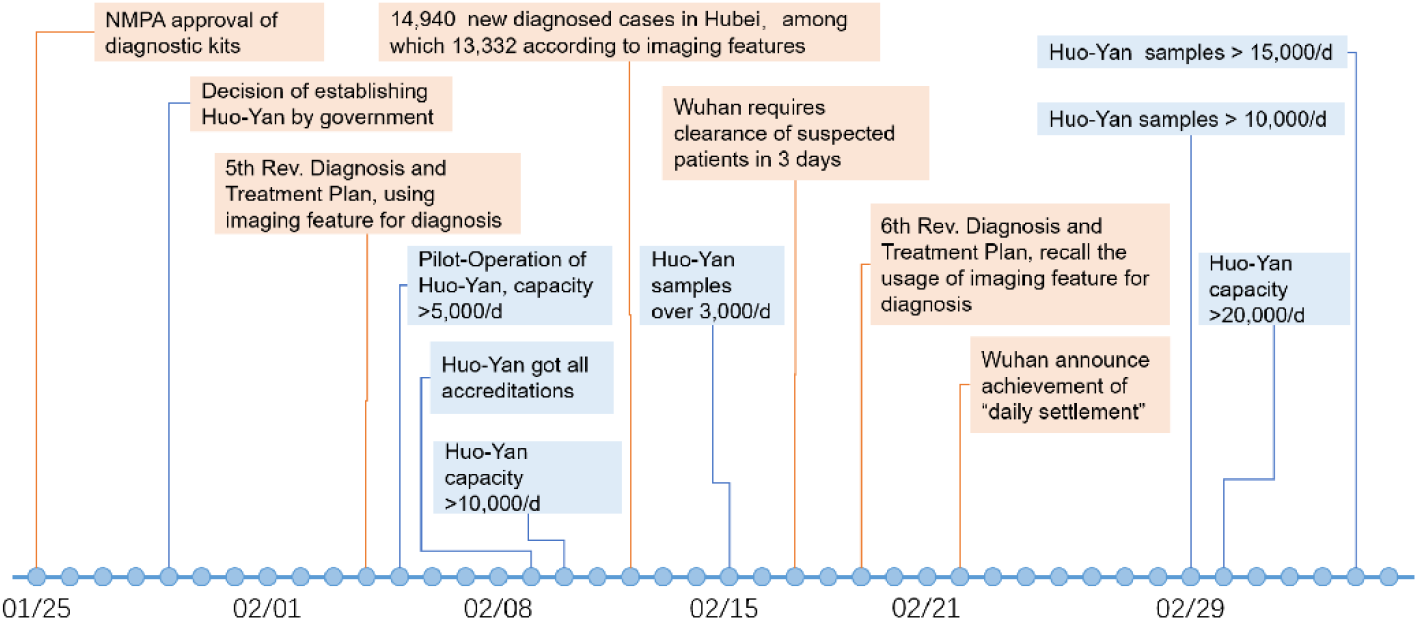
Timeline illustrating the establishment of Huo-Yan Laboratory as response to the insufficient nucleic acid testing during the epidemic.

Testing is the key to the prevention and control of infectious diseases, for only by identifying the infected can they be isolated and treated, as well as to stop the transmission. So far, there was no empirical data and numeric model to clarify the impact of standardized and large-scale clinical testing platform on the prevention and control of contagion. Here we present a novel differential model with non-linear transfer coefficients and competitive compartments to evaluate the trends of suspected cases under different nucleic acid testing capacities.

## Methods

### Data Source

The number of daily received samples and the maximum testing capacity of Huo-Yan Laboratory were taken into the model for the estimation of the testing (Figure 1) and online available (https://huoyan.bgi.com/#/). The data of suspected cases, diagnosed cases each day were acquired from the briefs released by the Health Commission of Hubei Province (http://wjw.hubei.gov.cn/) were used for validation of the model.

### Estimation of the number of nucleic acid testing carried out in Hubei Province

As the response to the rapid increment of suspected cases, the testing capacity of the hospitals, the local disease control and prevention institutions and the clinical testing laboratories in Hubei Province increased from c.a. 3,000 to over 30,000 tests per day. In the period of simulation (Feb 25^th^-March 6^rd^), Huo-Yan had finished over 163,000 testings by March 6^th^, with a team of 130 personals. Besides, the 20 teams of 83 personals sent by CCDCP together with local lab professionals and supporting personals, had finished 105,641 testings by the end of February. In Wuhan, the 23 most qualified hospitals could perform over 7,000 tests per day. The specifics of the testing carried out in Hubei Province were as follows:

- From January 19^th^, since the testing kits became available to hospitals and medical institutions, the daily testing capacity (TC) of Hubei Province was expected to be over 3,000;
- From Jan. 26^th^ to Feb. 11^th^, the daily testing capacity of clinical testing laboratories increased rapidly. Testing capacity of the Huo-Yan Laboratory, TCHY (t) increased to 10,000 per day on Feb 4^th^, and Huo-Yan accounted for 30%-45% of the testing in Hubei Province.xs
- From February 11^th^ to March 1^st^, TCHY (t) increased from 10,000 to 20,000 per day, delivered 40%-50% of the testing results in Hubei Province.

### Estimation of the total infected population of novel coronavirus and other pathogens

According to the modified SEIR model by *Yang et al* ^[1]^, after taken the whole province quarantine measures in Hubei, the infected cases decreased from 43,000 on Feb 25^th^ to 34,000 on Mar 6^th^. The suspected patents were usually with characteristics of fever and influenza-like illnesses (ILIs), and the existing epidemiological data showed an incremental trend of ILIs patients from 2015 to 2017 in Wuhan, along with annual ILIs prevalence of 4.5% in Wuhan ^[4]^. The ILIs cases in the 1^st^ quarter of each year accounted for about 20% (varying from 17% to 46%), therefore we estimated the annual total infected patients of ILIs other than coronavirus could be over 460,000 in the first quarter of 2020 in Hubei Province, which would lead to over 5,000 patients with similar symptom of COVID-19 per day.

### Model for predicting suspected cases

A novel model was used to illustrate the influence of testing capacity on the prevention and control of COVID-19 (Figure 3). Unlike the common dynamic model used only linear differential equations, this model applied the increasement of testing capacity into account. Since the quarantine measures in Hubei, the contact probability among people was reduced, which significantly reduced the possibility of large-scale transmission. Meanwhile, due to quarantine, people were more alert to fever and other symptoms, leading to more patients surged into the hospital and a continuous increasement suspected cases. The purpose of nucleic acid testing was to 1) identify patients with COVID-19 from the uninfected, and allow them to be hospitalized; 2) after the symptoms disappear, the inpatient with more than twice negative testing results (the interval must be more than 24 hours) could be discharged ^[3]^.

**Figure 2.**
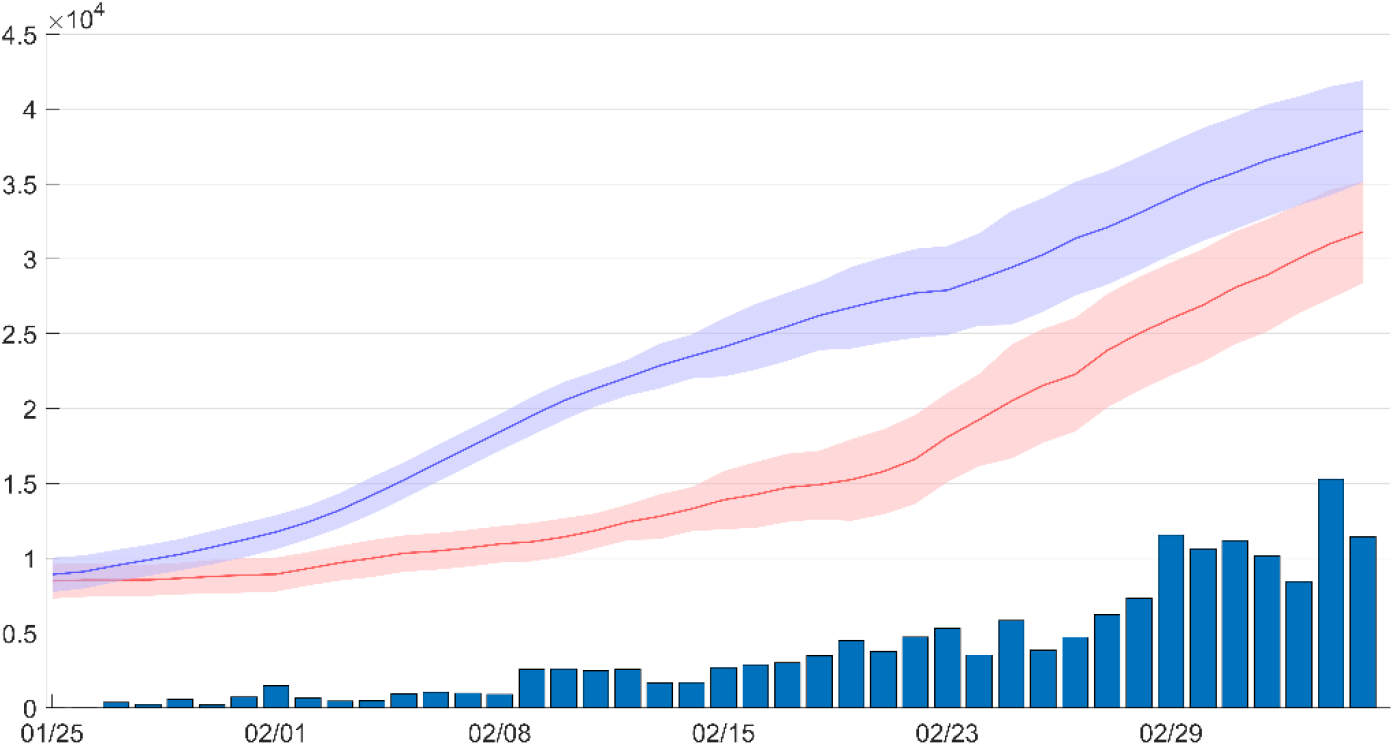
The trend of the nucleic acid testing performed in Hubei Province and by Huo-Yan Laboratory. The number of samples sent to Huo-Yan Laboratory and the corresponding delivered testing results (bar with solid line, blue). Estimated testing capacity of the Hubei Province (red line and the corresponding envelop) and the corresponding potential testing capacity (blue line and the corresponding envelop).

**Figure 3.**
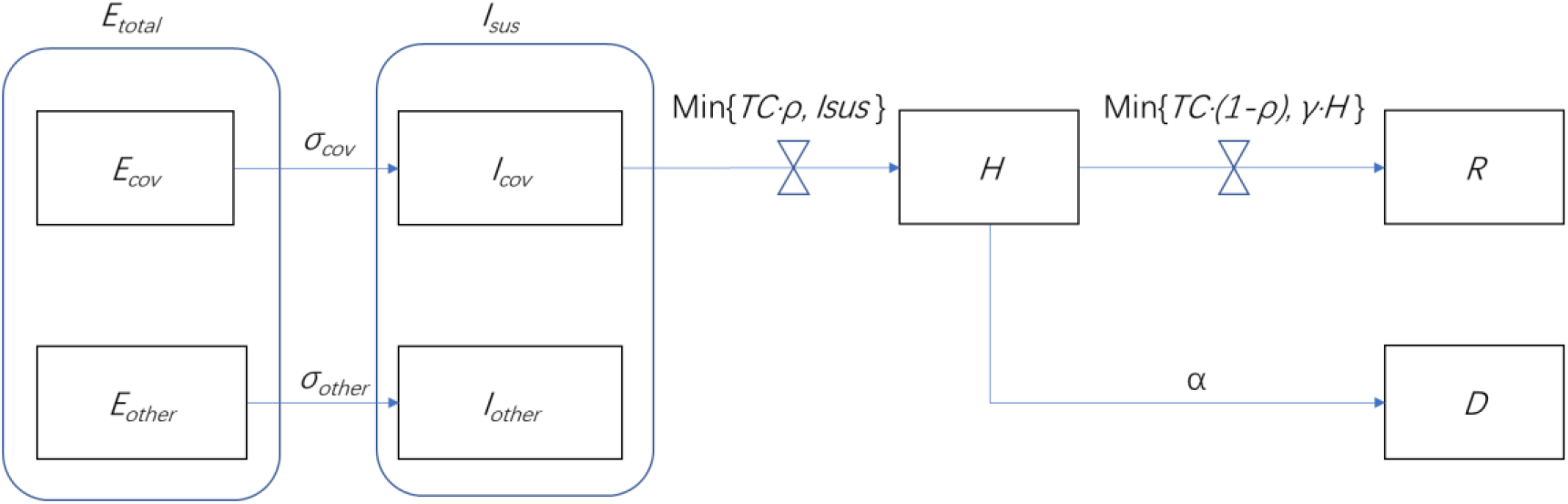
The competitive, non-linear epidemic model of hospitalization and discharge of the suspected patients. Unlike the common epidemiologic models based on the linear transfer functions and constant transfer coefficients, the novel model has transfer coefficient restricted by the testing capacity. *E*_*total*_ (t): the number of COVID-19 latent patients and other diseases in the region. *E*_*cov*_ (t): the latent patients of COVID-19 in the province, including the asymptomatic population. *E*_*other*_ (t): the latent patients that is not infected by the novel coronavirus. *σ*_*cov*_ : incubation rate. Generally, the reciprocal of the disease cycle is taken (1/7 day). *σ*_*other*_ : virtual incubation rate of other diseases that leads to symptom of suspected patients. (1 day). *I*_*sus*_ (t): the number of suspected cases of COVID-19 in the whole province. *I*_*cov*_ (t): the number of patients with novel coronavirus as suspected patients in the whole province. *I*_*other*_ (t): the number of patients of other diseases as suspected patients of COVID-19 in the whole province. *ρ* : the ratio of the test used for the diagnosis of the COVID-19 in the total nucleic acid tests. *TC*(t) : the testing capacity. *NTD*(t) : the number of tests used for diagnosis suspected patients. *NTR*(t) : the number of tests used for the discharge of the patients. *D*(t) : the cumulative number of deaths caused by COVID-19. *R*(t) : the cumulative number of discharged patients. *γ* : the probability of recovery, generally taking the reciprocal of 20 days. *α*: the mortality rate of COVID-19.

The conversion efficiency from suspected to hospital admission depended on the testing capacity (*TC*(t)), the number of existing and newly discovered the suspected cases, however there was a bottleneck of nucleic acid testing. As soon as the daily testing capacity was greater than the existing suspected plus the newly suspected of the day, the “daily settlement” of suspected cases could be achieved.

The differential equation derived from the following models:

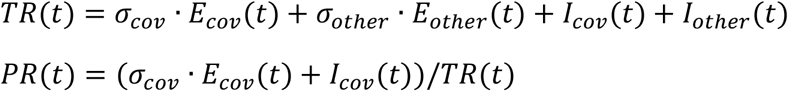

|  |  |  |
| --- | --- | --- |
| | $NTD(t) = \rho \cdot TC(t);$<br>$NTR(t) = 0$ | $when \rho \cdot TC(t) < TR(t)$ |
| | $NTD(t) = I_{sus}(t) + \sigma_{cov} \cdot E_{cov}(t);$ | $when TR(t) < \rho \cdot TC(t)$ |
| $\left\{ \begin{array}{l} \\ \\ \\ \end{array} \right.$ | $NTR(t) = (1 - \rho) \cdot TC(t)$ | *and* $TC(t) < (TR(t) + H(t))$ |
| | $NTD(t) = I_{sus}(t) + \sigma_{cov} \cdot E_{cov}(t);$<br>$NTR(t) = H(t)$ | *when* $TC(t) > (TR(t) + H(t))$ |

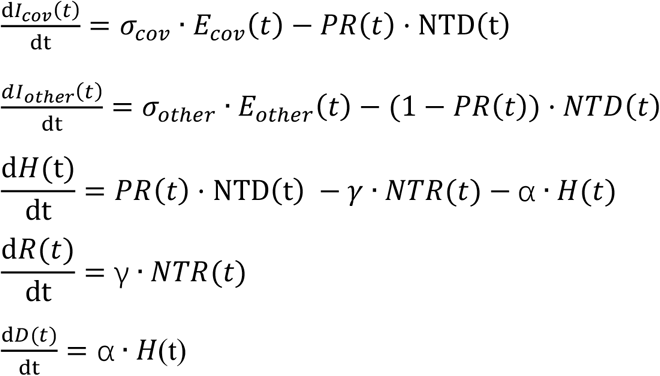

The parameters in the model were as follows:

*TC*(t): the testing capacity.

*TR*(t): the testing requirement from existing and newly discovered suspected patients.

*PR*(t): the positive ratio of the tests for diagnosis (*NTR*(t)).

*E*_*total*_ (t): the number of COVID-19 latent patients and other diseases in the province.

*E*_*cov*_ (t): the latent patients of COVID-19 in the province, including the asymptomatic population.

*E*_*other*_ (t): the latent patients that were not infected by the novel coronavirus.

*σ*_*cov*_: incubation rate. Generally, the reciprocal of the disease cycle was taken (1/7).

*I*_*sus*_ (t): the number of suspected cases of COVID-19 in the whole province.

*I*_*cov*_ (t): the number of patients with novel coronavirus as suspected patients in the whole province.

*I*_*other*_ (t): the number of patients of other diseases as suspected cases of COVID-19 in the whole province.

*ρ*: the rate of the test used for the diagnosis of the COVID-19 in the total nucleic acid tests.

*NTD*(t): the number of tests used for diagnosis suspected cases.

*NTR*(t): the number of tests used for the discharge of the cases.

*D*(t): the cumulative number of deaths caused by COVID-19.

*R*(t): the cumulative number of discharged patients.

*γ*: the probability of recovery, generally taking the reciprocal of 20 days.

*α*: the mortality rate, which is 0.0035.

## Results

The simulation result corresponded well with the trend of suspected cases by Health Commission of Hubei Province, and the positive rate of the tests per day was around 50%, also consistent with the positive rate data from Huo-Yan. The effect of increased testing capacity was significant, which was largely up to the government’s decision and the expansion of the hospitals and clinical testing laboratories (Figure 4).

**Figure 4.**
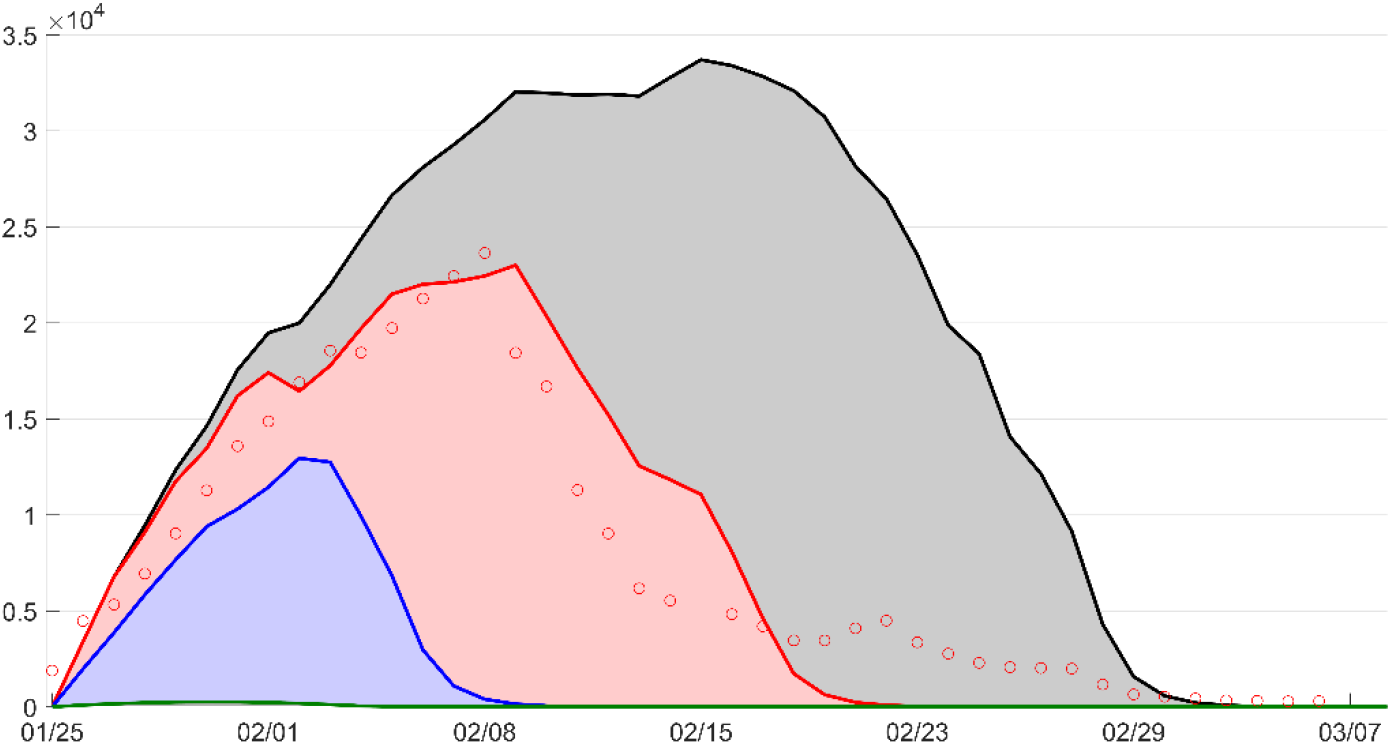
The simulation of the suspected patients under different testing capacity. The simulation results using the Huo-Yan factual operation data (line, red) corresponds with the open-access data from Health Commission of Hubei Province (circle, red). The simulated results without the Huo-Yan laboratory (line, black) and the result using 100% of the test capacity (blue). The ideal situation (line, green) would be achieved with a testing platform of enough high capacity. The area under curve depicts the number of isolated patients in term of rooms per person per day.

Due to the insufficient testing capacity at the beginning of the province-scale quarantine, the number of suspected cases rose to over 23,000, which became a “dammed lake” for delayed diagnosis and led to social panic.

If the testing capacity did not rapidly increased, the suspected cases could have reached a maximum of 33,700, resulting in a doubled isolation cost in term of room-days and ten thousands medical personals in addition to the over 40,000 medical workers and doctors which had been sent to Hubei Province. The diagnosis for over 30,000 suspected cases would be delayed, half of which are positive results and could cause further transmission. The turning point of the increment of suspected cases would be delayed for 6 days and the achievement of “daily settlement” would be realized at least 11 days later.

If the established testing capacity was fully used, over 22,800 suspected cases could be diagnosed on time rather than being delayed, and accordingly, the isolation cost could be reduced by at least 72%, the turning point of the increment of suspected cases could arrive one week earlier, and “daily settlement” could be realized 12 days in advance.

Under the ideal scenario, if Hubei Province was capable of carrying out more than 10,500 tests per day at the very beginning of the epidemic, there would be no “suspected cases” in the daily official COVID-19 epidemic report, but only the number of diagnosed cases, i.e., either positive or negative, because all of the suspected cases would be “settled” daily.

## Conclusions

Novel coronavirus related diseases have been officially defined as pandemic on March 11th 2020 by WHO. The quarantine of an entire district, city or a region could be adopted as part of the measures by the government. In Italy, more than 15 million people were placed in the country-based quarantine on March 8^th [5]^. Spain has announced it is placing its 47 million citizens under partial lockdown for 15 days. Hereby it would be worth determining the required testing capacity, referring Huo-Yan as an example in the public decision-making process. Timely and accurate clinical test is essential for identifying the infected, cutting off the transmission, and protecting the susceptible. The large-scale, precise, and reliable testing capacity is highly required to reduce the panic accompanied by the drastic quarantine measures.

To increase the testing capacity is a systematic project, among which the qualified laboratory spaces, the standardize of training medical panels, the supplement of the equipment, reagents, consumables and protective materials, and the automation of the testing procedure were of most importance. Here are the suggestions deduced from the simulation:

- The large-scale standardize testing platform and QC protocols were the premise of in the quarantine, which were the prerequisite for the diagnosis of suspected cases, isolate infectious patients, release isolation of convalescent and uninfected healthy population, and also the screening of key communities and groups. The practice from the centered platform could be summarized and replicated to other laboratories. The quality of the diagnostic kits and the accuracy, timeliness, safeties of the laboratories must be constantly compared and inspected. Unstandardized testing process would cause inconsistency in testing results and led to distrust on the testing results.
- Encourage the laboratories to increase testing capacities and keep continues delivery results at the same time. During emergencies, any changes in the testing process could cause samples accumulation, and the best solution could be continues applying new knowledge, know-hows in small scale and quickly replicate to the whole testing assembly line. This principle works in deploying new laboratory spaces, automation equipment, SOP and etc.
- Keep the outsourcing clinical testing laboratory the same priority and responsibility as the in-house laboratories. Despite of the high efficiency of the outsourcing laboratory, some hospitals are not willing to perform the outsourcing diagnostic tests just because they regard the risk of inaccurate testing results of the outsourcing laboratories “incontrollable”.
- Central planning of the diagnostic testing and comprehensive tracking of the potential testing capability to achieve “daily settlement”. Once there are standardize testing capacity, arrange the samples to fill the excess capacity immediately.
- Sufficiently supplement the sampling kits and the corresponding trainings to the medical panels. Other issues that require training on the sampling process including the barcoding and information input of the samples, and the inactivation of the pathogen before testing.
- Large-scale and standardized clinical laboratory should be regarded as infrastructure for both common time and emergencies against contagion, and should be put into use as early as possible in any epidemic. A good estimation of the testing capacity for nucleic acid testing of COVID-19 could be over 10,500 samples per day for a region of 60 million population with over 43,000 infected patients.

With Huo-Yan Laboratory as a reference model, combining with high-throughput sequencing, nucleic acid detection, immunoprotein analysis and other large-scale standardized and automated analysis methods, we can build infrastructure in the field of public health against the pandemic, so that large and small cities could have their own detection capabilities of 100 to 1,000 or 10,000 people when facing various epidemics, we can take it easy to ensure our life safety, biological safety and economic safety.

## Disclaimer

Huo-Yan is an ad-hoc COVID-19 clinical testing infrastructure owned by the Wuhan East Lake High-tech Development Zone. BGI-Wuhan operates the laboratory, BGI PathoGenesis Pharmaceutical Technology provides technical support for the whole laboratory. This work is to serve as an empirical reference to regions where COVID-19 needs to be prevented and controlled as it is now spreading globally. All opinions expressed are those of the authors and do not necessarily reflect the views of the Hubei provincial government.

## Data Availability

Some or all data, models, or code generated or used during the study are available from the corresponding author by request.

## Acknowledgement

Wang Jian supervised the whole work, Qing Xie generated the simulation model, Qing Xie, Jing Wang and Jianling You wrote the manuscript. We thank Dr. En Bo for his technical support.

## Notes

### Competing Interest Statement

The authors have declared no competing interest.

### Funding Statement

no external funding was received

